# Assessment of the Efficacy and Safety of Two Albendazole Regimens for the treatment of hypermicrofilaraemic loiasis in adult populations in Woleu-Ntem Province, Gabon: a Phase IIb single-blind randomised controlled trial

**DOI:** 10.1101/2025.05.22.25328128

**Authors:** Noé Patrick M’Bondoukwé, Luccheri Ndong Akomezoghe, Bridy Chesli Moutombi Ditombi, Jacques Mari Ndong Ngomo, Hadry Roger Sibi Matotou, Ginette Severine Zang Ondo, Valentin Migueba, Coella Joyce Mihindou, Bedrich Pongui Ngondza, Christian Mayandza, Héléna Kono, Dimitri Hardrin Moussavou Mabicka, Charleine Manomba, Reinne Moutongo, Luice Aurtin Joel James, Denise Patricia Mawili Mboumba, Marielle Karine Bouyou Akotet

**Author notes:** Corresponding author: (NPM).

## Abstract

**Background:** *Loa (L.) loa* hypermicrofilaraemia (>8,000 mf/mL) increases the risk of severe adverse events during mass ivermectin administration for onchocerciasis control. Albendazole has been proposed as a potential alternative for reducing microfilariaemia prior to treatment.

**Methodology and principal findings:** This prospective study was conducted in northern Gabon from November 2021 to April 2022. Individuals infected with *L. loa* were screened and allocated into three groups: two treatment arms receiving 400 mg or 800 mg of albendazole daily for 30 days among hypermicrofilaremic participants, and a control group with microfilaraemia <8,000 mf/mL. Clinical symptoms and parasitological data were collected on Days 0, 2, 7, 14, and 30. A total of 70 participants were enrolled: 16 in the 400 mg group, 16 in the 800 mg group, and 38 in the control group. Itching was the most common adverse event. By Day 30, no participants in the control group exhibited clinical symptoms. Microfilariaemia significantly decreased in all groups (p< 0.01). After 30 days, over 70.0% of patients treated with albendazole had microfilaraemia ≤8,000 mf/mL. There was no significant difference in efficacy between the two albendazole regimens.

**Conclusions/Significance:** Daily administration of 400 mg albendazole for 30 days effectively reduces microfilarial loads in patients with *L. loa* hypermicrofilaraemia and is well tolerated. This pre-treatment regimen may help mitigate the risk of adverse events associated with ivermectin administration. Further research is needed to evaluate the long-term persistence of microfilarial suppression.

**Author Summary:** Loiasis is a parasitic disease caused by the *Loa loa* worm and transmitted by the bite of a fly called *Chrysops*. People living in Central Africa can carry thousands of these microscopic worms in their blood. However, treating this disease is complicated: when the number of parasites in the blood is too high, giving the usual treatments (like ivermectin) can lead to dangerous side effects, including coma or even death. This study tested whether albendazole, a common anti-parasitic drug, could safely reduce the number of worms in heavily infected patients in rural Gabon. We compared two doses—400 mg and 800 mg taken daily for 30 days—and found that both doses were effective in lowering parasite levels below the danger threshold. The lower dose worked just as well and was better tolerated, making it more suitable for use in large-scale treatment campaigns. Our findings suggest that a 400 mg daily regimen of albendazole may help prepare patients for safer treatment with stronger medications and could improve control of this neglected tropical disease in hard-to-reach communities.

## Introduction

In areas where onchocerciasis is co-endemic with *Loa* (*L.*) *loa*, the mass drug administration (MDA) strategy using ivermectin (IVM) for onchocerciasis control is hampered by the presence of hypermicrofilaraemic individuals carrying more than 8,000 microfilariae (mf) per mL of blood [1]. These individuals are at higher risk of developing severe adverse reactions (SARs) following IVM administration. Moreover, the severity of these adverse effects increases with parasitic load: in individuals with microfilaraemia exceeding 30,000 mf/mL, the risk increases up to 200-fold, and in those with more than 50,000 mf/mL, up to 1,000-fold, potentially resulting in fatal outcomes [2,3]. SARs are common in nearly 10% of the exposed population in regions with high *L. loa* endemicity, such as in Gabon, where both filarial infections are prevalent. The prevalence of *L. loa* microfilaraemia exceeds 40.0% in some villages within onchocerciasis transmission areas (PNLMP).

Currently, no safe and effective treatment is recommended for hypermicrofilaraemic loiasis. Consequently, patients with such high parasitic densities are often left untreated when alternative non-pharmacological strategies are not available. In the “test and treat” strategy, the entire population is screened to identify hypermicrofilaraemic individuals; those with high microfilaraemia are excluded from mass IVM administration, thereby maintaining the reservoir of *L. loa* within the community [4].

A treatment allowing a reduction of hypermicrofilaraemia below the 8,000 mf/mL threshold for a sufficiently long period would enable subsequent IVM administration and may lead to the control and elimination of both onchocerciasis and lymphatic filariasis (LF) in areas co-endemic with *L. loa*. The World Health Organization (WHO) has suggested semiannual treatment with albendazole (ALB), complemented by vector control measures, as a potential strategy for LF elimination in *L. loa* transmission areas [5]. This molecule could also be used to reduce hypermicrofilaraemia prior to mass IVM administration (IVM MDA) in well-defined onchocerciasis foci. For more than 30 years, albendazole has been considered an alternative to diethylcarbamazine (DEC) or IVM for the curative treatment of *L. loa* [6]. It acts on microfilariae and is associated with a lower risk of post-therapeutic SARs compared to DEC or IVM [7]. Recently, it has been reported that administering 2 to 6 doses of 800 mg albendazole, at two-month intervals, can reduce *L. loa* microfilaraemia by at least 50.0% and potentially maintain parasitaemia below 8,000 mf/mL for at least 4 months [4]. However, there remains controversy regarding the microfilaricidal effect of albendazole, and no consensus exists on the optimal therapeutic regimens or treatment duration for hypermicrofilaraemic *L. loa*.

In Gabon, DEC is no longer available. There is a need to provide evidence-based information to support national and international recommendations for managing hypermicrofilaraemic *L. loa* using approaches that exclude both IVM and DEC. Moreover, it is important to use effective molecules for parasitic diseases in the same geographical areas, as recommended by the NTD Roadmap[8]. The DPMTM serves as the reference centre for medical parasitology and maintains one of the largest cohorts of *L. loa* patients. Weekly consultations, along with clinical management for filariasis patients, are conducted at this centre. Albendazole has been routinely used for the curative treatment of *L. loa* since the interruption of DEC and the Mectizan programme over the past 10 years, based on clinical data [9,10]. Recently, DPMTM reported that administration of 400 mg albendazole showed a reduction rate of more than 80.0% in microfilaraemia in a clinical study conducted in Gabon among patients with low microfilaraemia [11]. It is also necessary to demonstrate whether this effective protocol can be applied to hypermicrofilaraemic participants to enable their eligibility for IVM administration.

Thus, the present study aims to identify an optimal treatment regimen for reducing *L. loa* hypermicrofilaraemia in patients living in hyperendemic areas of northern Gabon.

## Material and Methods

### Study Area

This study was conducted in Gabon, a Central African country bordered to the north-west by Equatorial Guinea, to the north by Cameroon, to the east and south by the Republic of the Congo, and to the west by the Atlantic Ocean (Fig 1). In 2021, its population was estimated at 2.1 million inhabitants. Located in the heart of Africa and straddling the Equator, Gabon is administratively divided into nine provinces, each with its own geographical and cultural characteristics.

**Fig 1.**
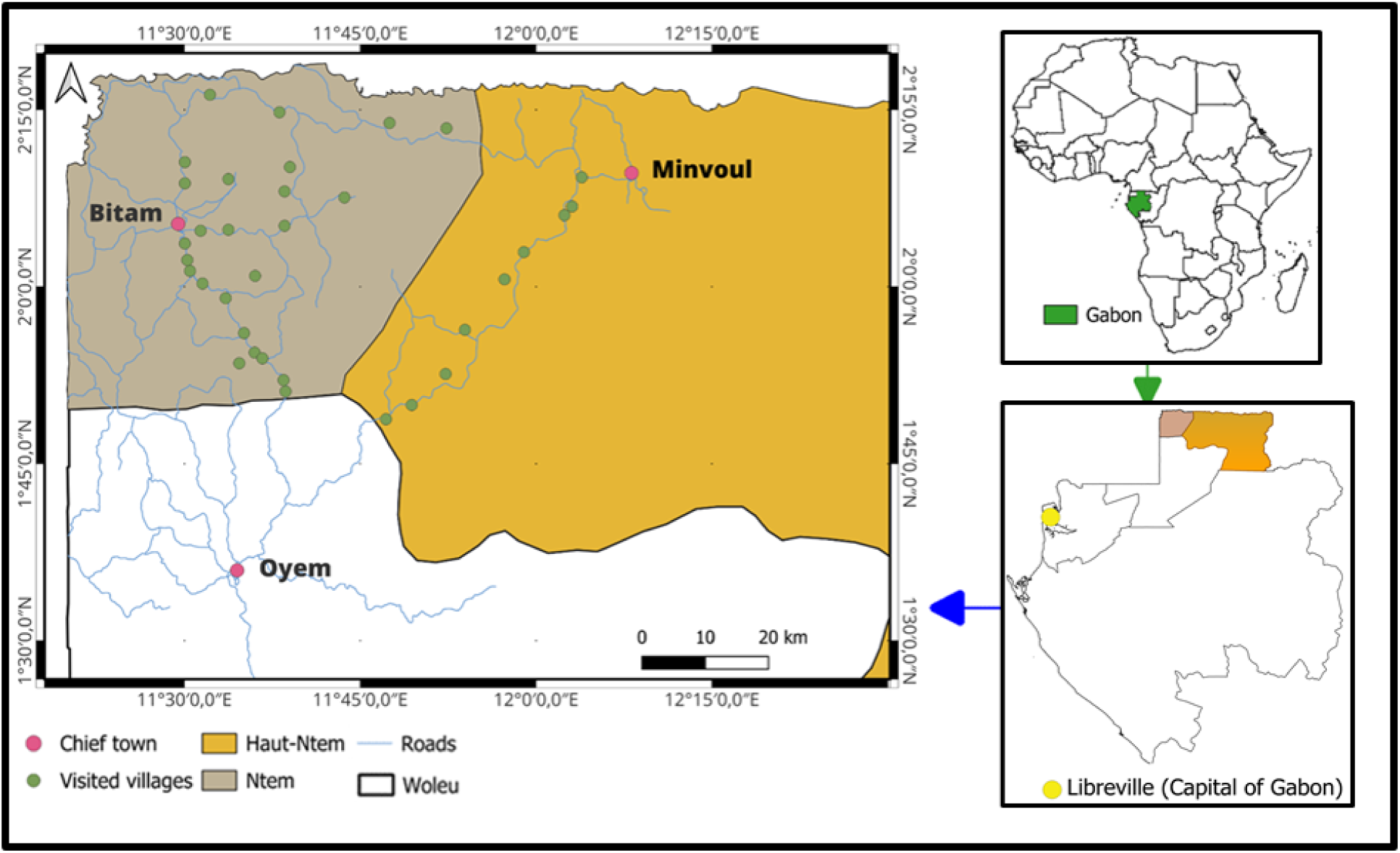
Study areas. The green plots corresponds to villages where participants of the clinical trial were screened and/or recruited.

The country is traversed by several major rivers, most notably the Ogooué River, Gabon’s principal watercourse, which spans approximately 1,200 km, draining nearly 80% of the national territory. Among its major tributaries are the Ivindo River, which extends for 500 km, and the Ngounié River, which runs for 300 km.

Gabon stretches about 800 km in length and varies from 20 to 300 km in width, with approximately 80% of its area covered by dense, tropical rainforest. This vast forested region extends from west to east, beginning with a coastal basin characterised by grassland forests, progressing into the interior’s plateau forests in the north-east, and including a broad mountainous and forested belt of 60 to 100 km running parallel to the coast. Isolated savannah and steppe areas are found in the south and south-east of Gabon, contributing to the country’s rich and diverse ecosystems [12].

### Study Site

This study was conducted among the rural population of Woleu-Ntem, specifically in the departments of Ntem and Haut-Ntem. The region has a population of approximately 155,000 inhabitants, with a density of about 4 inhabitants per km². Woleu-Ntem is a province located in northern Gabon. It covers an area of 38,465 km², and its capital city is Oyem.

The province has a hot and humid equatorial climate with heavy rainfall. It is predominantly covered by secondary forests and boasts a rich and diverse fauna. The prevalence of *L. loa* in the region is estimated at 20.2% [12]. Woleu-Ntem is a relatively under-industrialised area known for producing cocoa, coffee, and rubber. Subsistence farming and hunting in the forest still constitute significant parts of the local diet. The economy of the province revolves around agricultural produce. There are strong ties with Equatorial Guinea and southern Cameroon, partly driven by shared Fang ethnic heritage among the majority of the population on both sides of the border.

Villages around Bitam (Ntem) and Minvoul (Haut-Ntem) were selected for this clinical study, where all participants lead a rural lifestyle that exposes the population to the *Chrysops* sp. vector of *L. loa* (Fig 1).

### Type and Study Period

This is a Phase IIb randomised, single-blind clinical trial conducted from November 2021 to April 2022.

### Study Population

#### Inclusion Criteria

The inclusion criteria were as follows:

- Age between 18 and 75 years;
- Weigh less than 90.1 kg;
- *L. loa* microfilaraemia (≥8,000 mf/mL for the treatment group and <8,000 mf/mL for the comparison group);
- Signing an informed consent form;
- Agreement to comply with the study procedures, including blood sample collection;
- Residing in or being in the village during the clinical trial period.

#### Non-Inclusion Criteria

The non-inclusion criteria were as follows:

- Having declared chronic diseases such as HIV/AIDS, acute or chronic hepatitis, cancer, diabetes, chronic heart disease, or renal disease;
- Taking anthelmintics such as diethylcarbamazine (DEC), suramin, ivermectin, mebendazole, or albendazole within four (4) weeks prior to screening;
- Being allergic to benzimidazoles;
- Being pregnant or breastfeeding.

#### Exclusion Criteria

The exclusion criteria were as follows:

- Withdrawal of consent to participate in the study;
- Worsening of symptoms or the appearance of severity criteria related to *L. loa* or another disease;
- An increase in microfilaraemia of more than 50% compared to the initial value.

### Sample Size Calculation

The pwr.anova.test function in R version 4.4.4 was used to determine the sample size for clinical trials where the primary endpoint is a continuous variable that does not necessarily follow a normal distribution. Additionally, the number of treatment arms was set at three. *L. loa* microfilaraemia was the primary endpoint for both hypermicrofilaraemic participants and the control group with microfilaraemia below 8,000 mf/mL.

The following formula, based on Cohen’s guidelines, was applied, where “k” denotes the number of comparison groups and “f” the effect size, with Cohen’s f values of 0.1, 0.25, and >0.4 representing small, medium, and large effect sizes, respectively.

Considering three treatment arms (k = 3), a large effect size (f = 0.6), an alpha risk of 5%, and a statistical power of 80%, a minimum of 10 patients completing treatment in each group would be required to ensure sufficient power to validate the results.

### Study design

#### Study Drugs

The investigational product was Ubigen® albendazole 400 mg (UBITHERA PHARMA PVVT. Ltd.). The 400 mg tablets were administered orally under direct supervision after a fatty meal, which is essential to optimise absorption and efficacy.

#### Control group

Patients in this group, with microfilaraemia below 8,000 mf/mL, received a single daily dose of 400 mg (one 400 mg tablet) for 30 days. Additionally, they received a 10 mg cetirizine tablet daily for 7 days. This procedure was carried out under the supervision of a designated community worker within the village to ensure adequate follow-up.

#### Albendazole 400 mg group

Patients in this group, with microfilaraemia ≥8,000 mf/mL, received a single daily dose of 400 mg (one 400 mg tablet) for 30 days with the procedure supervised by the village community worker.

#### Albendazole 800 mg group

Patients in this group, also with microfilaraemia ≥8,000 mf/mL, received a daily dose of 800 mg, which corresponds to two 400 mg tablets administered as a single dose for 30 days. They similarly received a 10 mg cetirizine tablet daily for 7 days, all under the supervision of the village community workers.

#### Safety Parameters

Safety variables included clinical signs and symptoms related to *L. loa* infection, both common and uncommon, such as myalgia/arthralgia, headaches, asthenia, pruritus, transient nerve paralysis, subcutaneous or ocular circulation related to adult worms, vision disorders, and Calabar swelling. The frequency of adverse events (AEs) up to one month after the first dose of albendazole was monitored and classified according to type, severity, and relationship to the investigational drug, as assessed by the investigators.

Severity was classified as:

- *Mild*: The event is tolerated by the participant, causing minimal discomfort and not interfering with daily activities;
- *Moderate*: The event is uncomfortable enough to interfere with daily activities;
- *Severe*: The event prevents normal daily activities;
- *Not applicable*: Events for which the intensity cannot be meaningfully assessed.

Serious adverse events (SAEs) were evaluated according to the ICH definition: any untoward medical occurrence that, at any dose, results in death, is life-threatening, requires hospitalisation or prolongs an existing hospitalisation, results in persistent or significant disability/incapacity, or is a congenital anomaly/birth defect, and is related to any dose of a drug or the doses normally used in humans [1].

#### Data Collection and Screening

A preliminary visit was conducted to inform local authorities about the study’s objectives. After obtaining informed consent from participants, sociodemographic and clinical data were collected, including age, gender, weight, height, and clinical manifestations attributable to *L. loa*. Subsequently, a 4 mL venous blood sample was collected in an EDTA tube for parasitological screening of *L. loa* via direct examination of 10 µL of blood.

#### Patients Randomisation

Taves’ covariate-adaptive randomisation was used to allocate participants across the different treatment arms [13]. The covariates included gender (male, female), age groups (18–64 years, over 64 years), and microfilaraemia strata (less than 8,000 mf/mL, 8,000–30,000 mf/mL, 30,001–50,000 mf/mL, and over 50,000 mf/mL). Patients were randomised according to a 2:1:1 ratio. The list of eligible participants by stratum was provided to an independent pharmacist, who was not involved in the study. The pharmacist was responsible for assigning the treatment indicated on the randomisation list based on the participant’s position on the eligible list for each stratum. Additionally, the pharmacist administered the first dose of treatment.

#### Patients Follow-Up

Following the administration of the first dose, ongoing treatment was supervised by focal points—community workers selected within the villages—or by the study team. The treatment was taken 15 to 30 minutes after a fatty meal (such as fatty rolls with approximately 15 g of butter). On scheduled visit days and in villages without a community worker, a mobile team comprising the study physician and/or a pre-trained field health worker responsible for treatment administration, identification, and reporting of *L. loa*-related symptoms, as well as detection, management, and notification of potential adverse events of albendazole, supervised the treatment. During these visits, venous blood samples in EDTA tubes were also collected to monitor microfilaraemia.

### Parasitological Diagnosis of *L. loa* Microfilaraemia

#### Direct Examination of 10 µL of Blood

A direct examination was performed to detect and quantify microfilariae, thereby assessing the presence and intensity of infection. This method is crucial for diagnosing and initiating treatment. The slide examination was conducted during the peak hours of the day following sample collection to maximise parasite motility, which is essential for species identification. Ten microlitres of blood were carefully placed on a microscope slide and covered with a coverslip. The sample was examined under a microscope (magnification ×10). Parasitaemia was expressed as the number of microfilariae per millilitre of blood (mf/mL), providing vital quantitative data for infection assessment.

#### Leucoconcentration of 4 mL of Blood

The concentration technique was performed according to the method described by Ho Thi Sang and Petithory (1964), which offers increased sensitivity for detecting microfilariae [14]. A volume of 4 mL of peripheral blood was centrifuged at 2000 rpm for 5 minutes. After centrifugation, the plasma was carefully removed using a pipette. Then, 2 mL of physiological saline (0.9% NaCl) and 1 mL of a 2% saponin solution were added to the red cell pellet, and the mixture was homogenised to ensure complete lysis of red blood cells. A 10-minute waiting period was observed to allow haemolysis. Following this, the tube was centrifuged again at 2000 rpm for 10 minutes, and the supernatant was discarded. The NaCl and saponin procedure was repeated before the final centrifugation. The resulting white leukocyte pellet was washed with 3 mL of 0.09% NaCl. Once free of debris, the leukocyte pellet was placed between a slide and a coverslip and examined under an optical microscope using a ×10 objective for locating microfilariae and a ×40 objective for identification and quantification.

### Ethical considerations

This study was conducted as part of the PHYLECOG project, financed by the EDTCP2 programme under reference TMA2019CDF-2730. The study protocol was reviewed and approved by the National Ethics Committee for Scientific Research (CNER) (Protocol No. 0053/2022/CNER/P/SG). Written informed consent was obtained from each participant after explaining the study to local and health authorities. The study was registered under the reference ISRCTN14889921.

### Statistical Analyses

Data processing was carried out using REDCap. All analyses were performed using R software version 4.4.2. The significance threshold was set at 5% for all tests. Categorical data, such as gender, symptoms, and adverse events, were presented as frequencies and percentages. The microfilaraemia reduction rate was calculated as a percentage using the following formula:

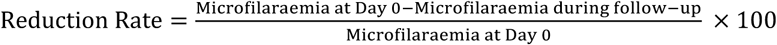

Percentage comparisons were made using the Chi-square test for reduction rates between visits namely the proportions of patients achieving the primary endpoint (microfilaraemia below 8,000 mf/mL) at different follow-up visits.

Survival analysis, typically Kaplan-Meier curves, was used to graphically depict the number of patients maintaining microfilaraemia below 8,000 mf/mL over time. The primary endpoint (reduction of microfilaraemia to below 8000 mf/mL) was analysed on both a per-protocol and an intention-to-treat basis.

## Results

### Patients

Among the 1,342 volunteers screened for *L. loa* microfilaraemia, 70 participants met all the inclusion criteria for the clinical trial. Of these, 38 had microfilaraemia below 8,000 mf/mL, and 32 had microfilaraemia above 8,000 mf/mL. Participants in the control group received 400 mg of albendazole (ALB) daily for 30 days; 16 participants with microfilaraemia at or above 8,000 mf/mL received 400 mg ALB daily for 30 days, and the remaining 16 received 800 mg ALB daily for 30 days (Fig 2).

**Fig 2.**
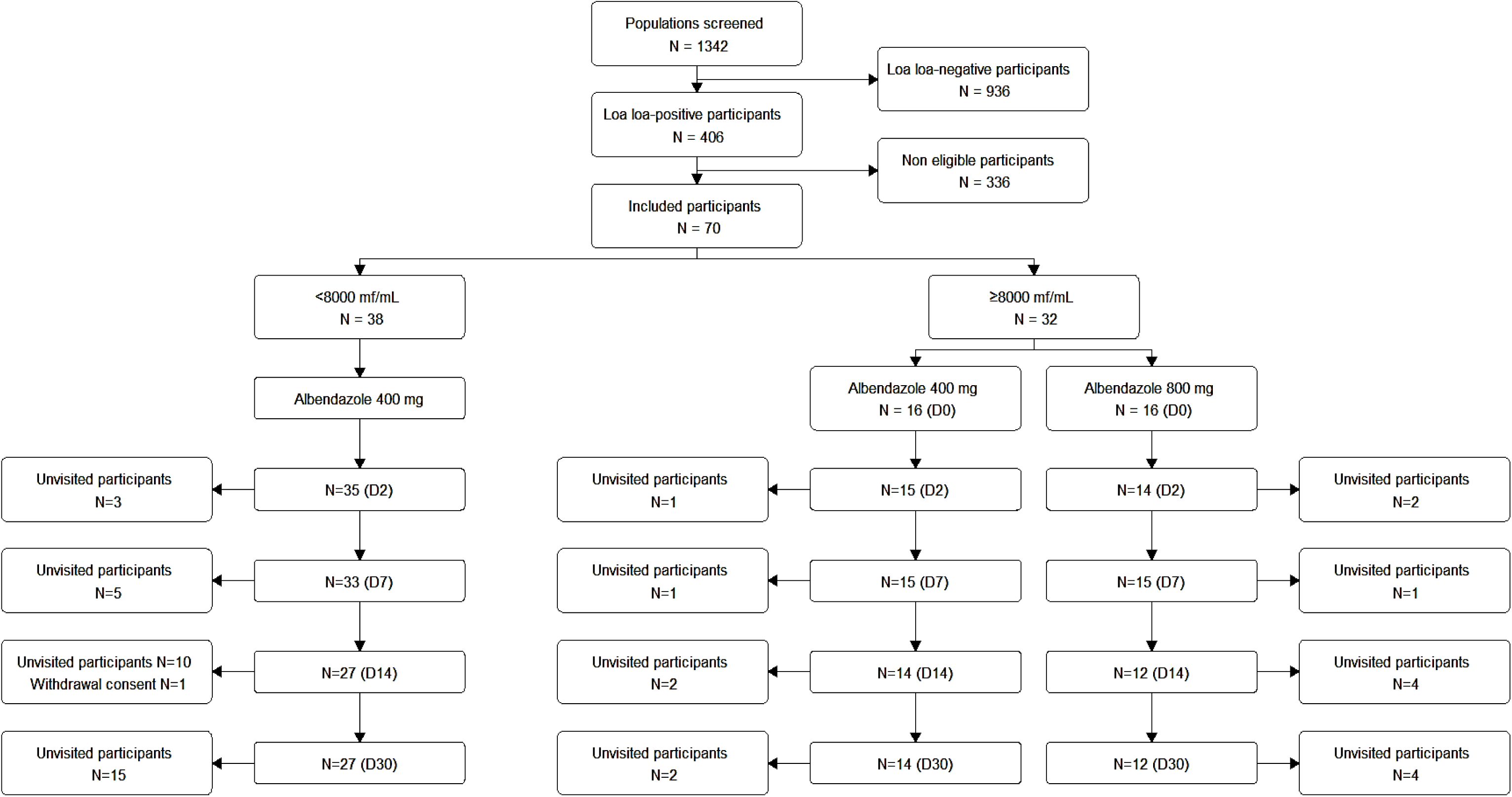
Study flowchart. Flow diagram showing the number of participants screened and enrolled in the clinical study. One participant in the control group withdrew consent. No participants were lost to follow-up, as the study design allowed visits at two non-consecutive time points.

Table 1 summarises the characteristics of the study population prior to treatment. There were no significant differences between the three treatment groups for variables such as sex, age, weight, height, and eosinophil count. Regarding the mean microfilarial load, it was comparable between the two treatment arms of hypermicrofilaraemic participants (p = 0.7).

**Table 1.** Descriptive characteristics of treatment group at the inclusion.

| **Variables** | **Control group (N= 38)** | **Albendazole 800 mg (N=16)** | **Albendazole 400 mg (N=16)** | ***p*-value** |
| --- | --- | --- | --- | --- |
| **Sex, n (%)** | 14 (36.8) | 4 (25.0) | 5 (31.2) | 0.7 |
| **Age in years, mean ± SD<sup>1</sup>** | 49.8 ± 12.4 | 52.2 ± 12.4 | 46.7 ± 12.8 | 0.4 |
| **Weight in Kg, mean ± SD** | 64.2 ± 10.4 | 60.2 ± 10.6 | 63.0 ± 10.1 | 0.8 |
| **Height in cm, mean ± SD** | 164.3 ± 8.4 | 167.6 ± 9.3 | 162.4 ± 7.0 | 0.1 |
| **Microfilaraemia in mf/mL, geometric mean** | 2258 | 17249 | 19178 | NA<sup>2</sup> |
| **Eosinophilia rate in %, mean ± SD** | 20.4 ± 3.0 | 15.1 ± 7.0 | 18.7 ± 5.0 | 0.7 |

**SD<sup>1</sup>** : Standard deviation
**NA<sup>2</sup>** : Not applicable for the three groups.

### Reduction in Microfilaraemia in Intention-to-Treat and Per-Protocol Analyses

Compared to the initial parasitaemia (Day 0), both the intention-to-treat (ITT) and per-protocol (PP) analyses demonstrated significant reductions in microfilarial loads across all study groups by Day 30 (p < 0.01). In the control group, the geometric mean decreased from 2,252 mf/mL to 258 mf/mL in the ITT analysis (−88.5%), and from 1,938 mf/mL to 231 mf/mL in the PP analysis (−88.1%).

For participants treated with 400 mg of albendazole, microfilaraemia decreased from 17,249 mf/mL to 4,177 mf/mL (−75.8%) in the ITT analysis, and from 18,191 mf/mL to 4,177 mf/mL (−77.0%) in the PP analysis. In the 800 mg albendazole group, the geometric mean decreased from 19,178 mf/mL to 1,987 mf/mL (−89.6%) in ITT, and from 17,991 mf/mL to 2,352 mf/mL (−86.9%) in PP (Fig 3A and 3B).

**Fig 3.**
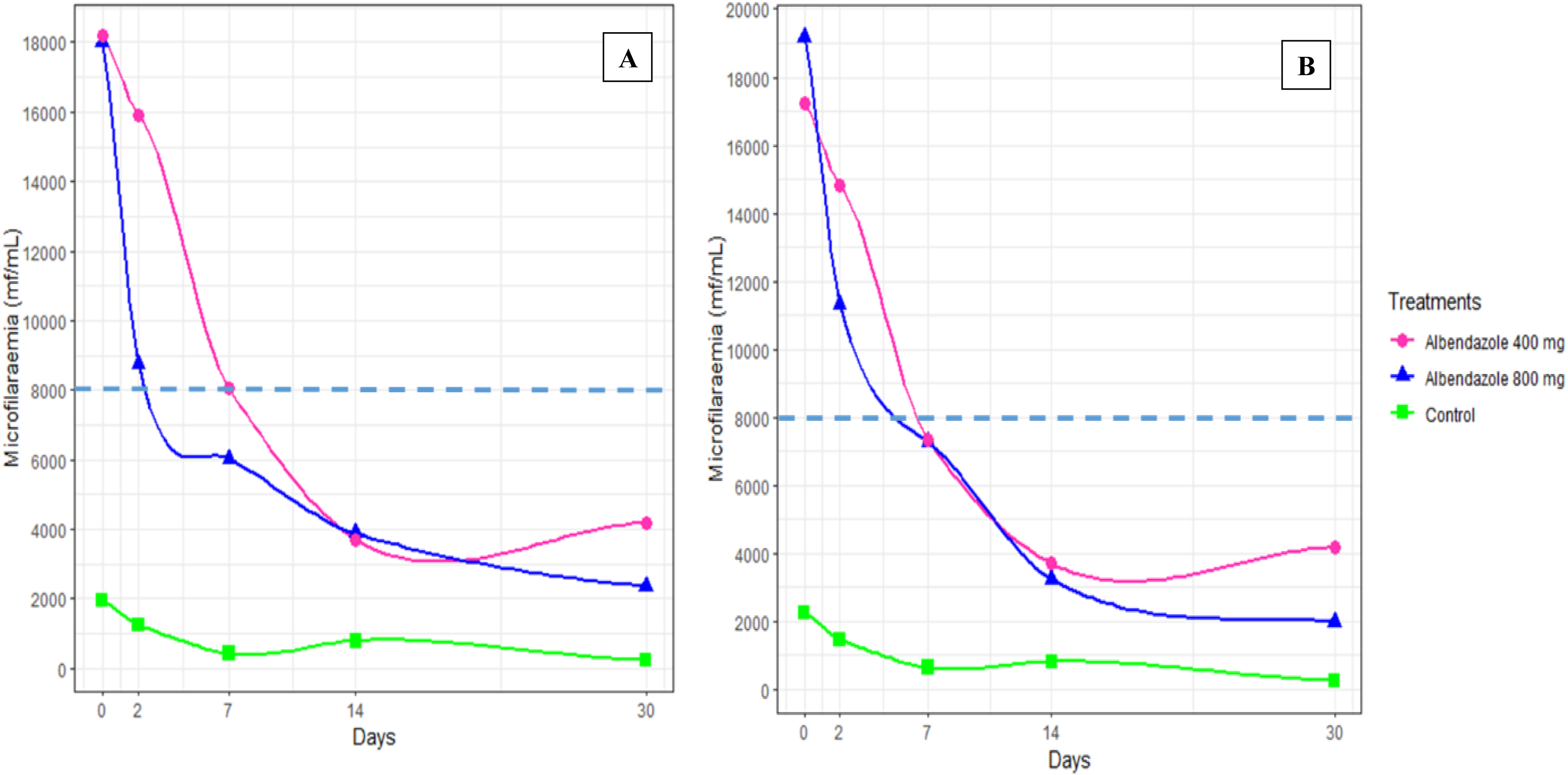
Evolution of microfilaraemia in the treatment and control groups from Day 0 to Day 30. Panel A shows the per-protocol analysis, while Panel B presents the intention-to-treat analysis.

Daily reduction trends were consistent between the two analyses. In the control group, reductions on Day 2 were 34.7% (ITT) and 36.1% (PP), increasing to 71.1% and 78.3% respectively by Day 7. In the 400 mg group, reductions on Day 2 were 14.0% (ITT) and 12.5% (PP), reaching 78.6% (ITT) and 79.7% (PP) by Day 14. The most pronounced reductions were observed in the 800 mg group: 40.9% (ITT) and 51.5% (PP) at Day 2, and 89.6% (ITT) and 86.9% (PP) by Day 30 (Fig 3A and 3B).

### Reduction in Microfilaraemia below 8000 mf/mL according to Intention-to-Treat and Per-Protocol Analyses

In both treatment groups, the geometric mean of microfilaraemia dropped below the 8,000 mf/mL threshold before Day 7 of treatment. However, this decrease occurred earlier in the 800 mg group, while patients receiving 400 mg still had a mean microfilaraemia of 8,055 mf/mL at Day 7 in the PP analysis (Fig 4A and 4B).

**Fig 4.**
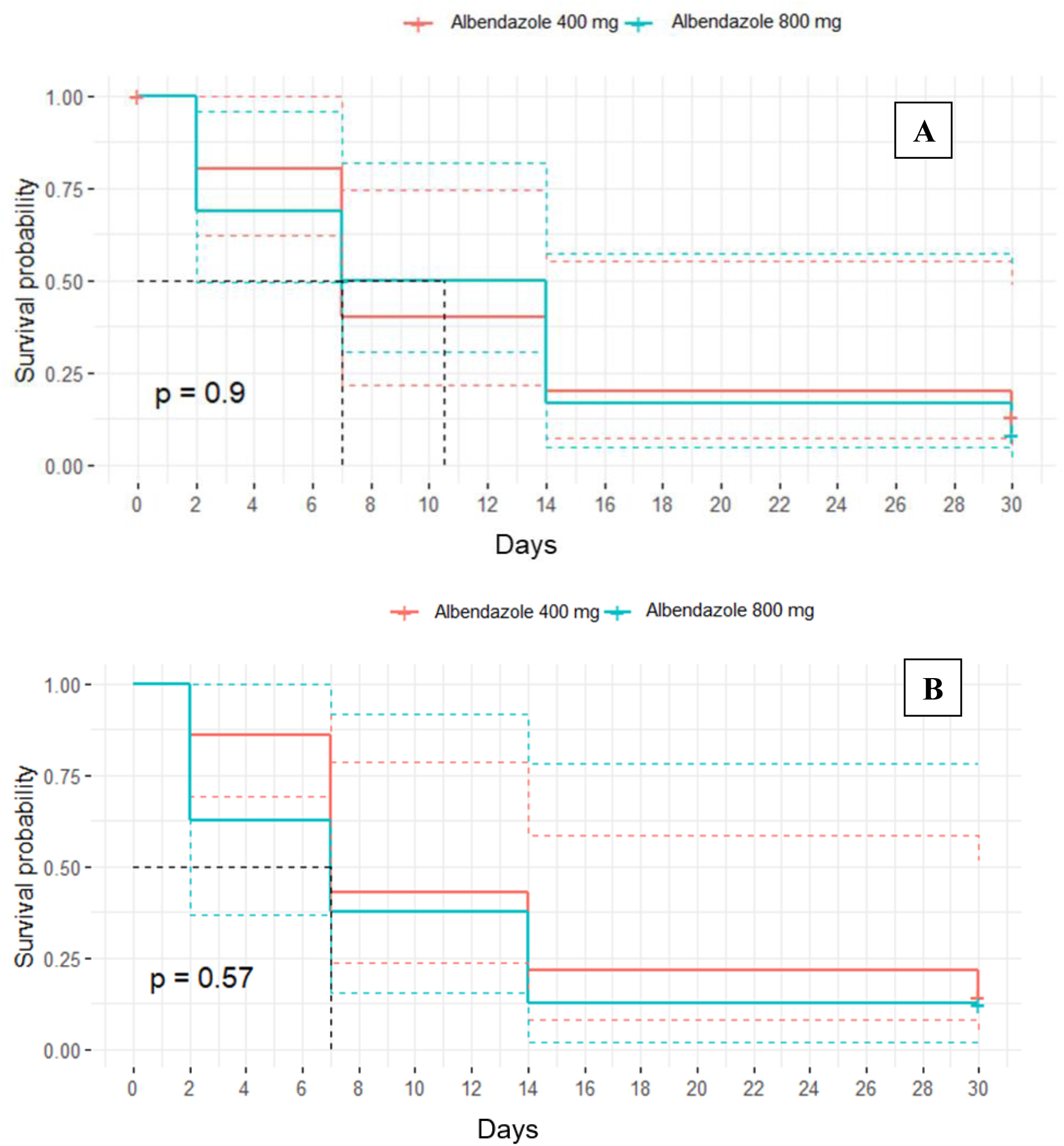
Survival analysis. This figure illustrates the proportion of hypermicrofilaraemic participants in the two treatment arms over time, in both the intention-to-treat (ITT) (A) and per-protocol (PP) (A) analyses.

The probability that a patient would have microfilaraemia above 8,000 mf/mL decreased progressively over the 30-day follow-up in both treatment arms. On Day 2, this probability was estimated at 0.80 [0.62; 1.0] for the 400 mg group and 0.68 [0.49; 0.96] for the 800 mg group in the ITT analysis, while in the PP analysis, it was 0.86 [0.69; 1.0] and 0.62 [0.36; 1.0], respectively. These correspond to reductions of approximately 14–20% with 400 mg and 32–38% with 800 mg.

By Day 7, the probability further decreased to 0.40 [0.21; 0.74] (400 mg) and 0.50 [0.31; 0.81] (800 mg) in the ITT analysis, and 0.43 [0.23; 0.78] and 0.37 [0.15; 0.92] in the PP analysis—indicating reductions ranging from 50.0% to 63.0%.

At Day 14, probabilities declined further to 0.20 [0.07; 0.55] and 0.17 [0.05; 0.57] for the 400 mg and 800 mg groups, respectively, in the ITT analysis, and 0.21 [0.08; 0.58] and 0.12 [0.02; 0.78] in the PP analysis—equivalent to reductions of approximately 79% to 88%.

Finally, at Day 30, the probability of remaining above the 8,000 mf/mL threshold was 0.13 [0.09; 0.48] in the ALB 400 mg group and 0.08 [0.01; 0.53] in the ALB 800 mg group in the ITT analysis, while the PP analysis reported 0.14 [0.04; 0.51] and 0.12 [0.02; 0.78], respectively. These correspond to reductions of approximately 86% to 92%.

Despite these encouraging trends, no statistically significant difference was observed between the two dosages. In terms of patient distribution, half of the participants had microfilaraemia below 8,000 mf/mL by Day 7 in both treatment groups; however, the ITT analysis suggested this occurred slightly earlier for the 400 mg group (Day 7) and later for the 800 mg group (Day 11).

### Evolution of symptoms

At baseline, 15.7% of patients (n = 11/70) presented with at least one symptom. The most common symptoms were ocular migration of a worm (26.7%, n = 6/11) and pruritus (24.1%, n = 5/11). The median number of symptoms was 2 [1–2] (Table 2).

**Table 2.**
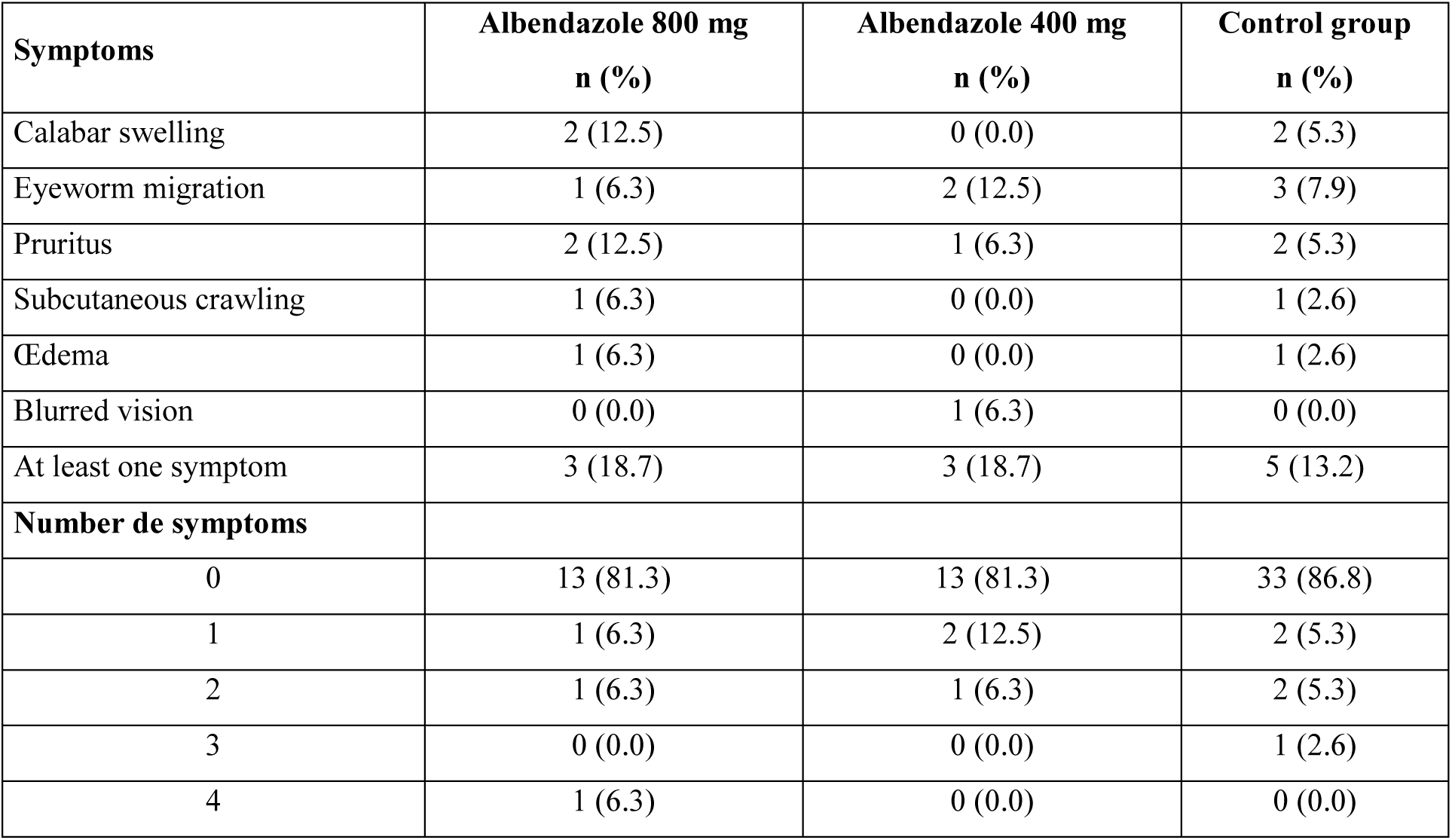
Proportions of symptoms at the inclusion Day 0.

In the control group, 13.2% of patients (n = 5/38) exhibited at least one symptom, with the most frequent being sub-conjunctival migration of the adult worm, observed in 7.9% of patients. In the group receiving 400 mg of albendazole, 18.7% (n = 3/16) presented with symptoms, most notably sub-conjunctival migration, seen in 12.5% of patients. Among those treated with 800 mg of albendazole, 18.7% (n = 3/16) exhibited at least one symptom; the most common were pruritus and Calabar swelling (Table 2).

In all three groups, although the frequency of other symptoms tended to decrease throughout the course of treatment, pruritus, on the contrary, tended to increase before disappearing by the end of the 30-day treatment period (Fig 5). With the exception of the group that received 400 mg of albendazole, in which 14.3% and 7.1% of patients reported pruritus and blurred vision respectively after 30 days of treatment, no symptoms were reported in any of the treatment groups at the end of the 30-day period (Fig 5).

**Fig 5.**
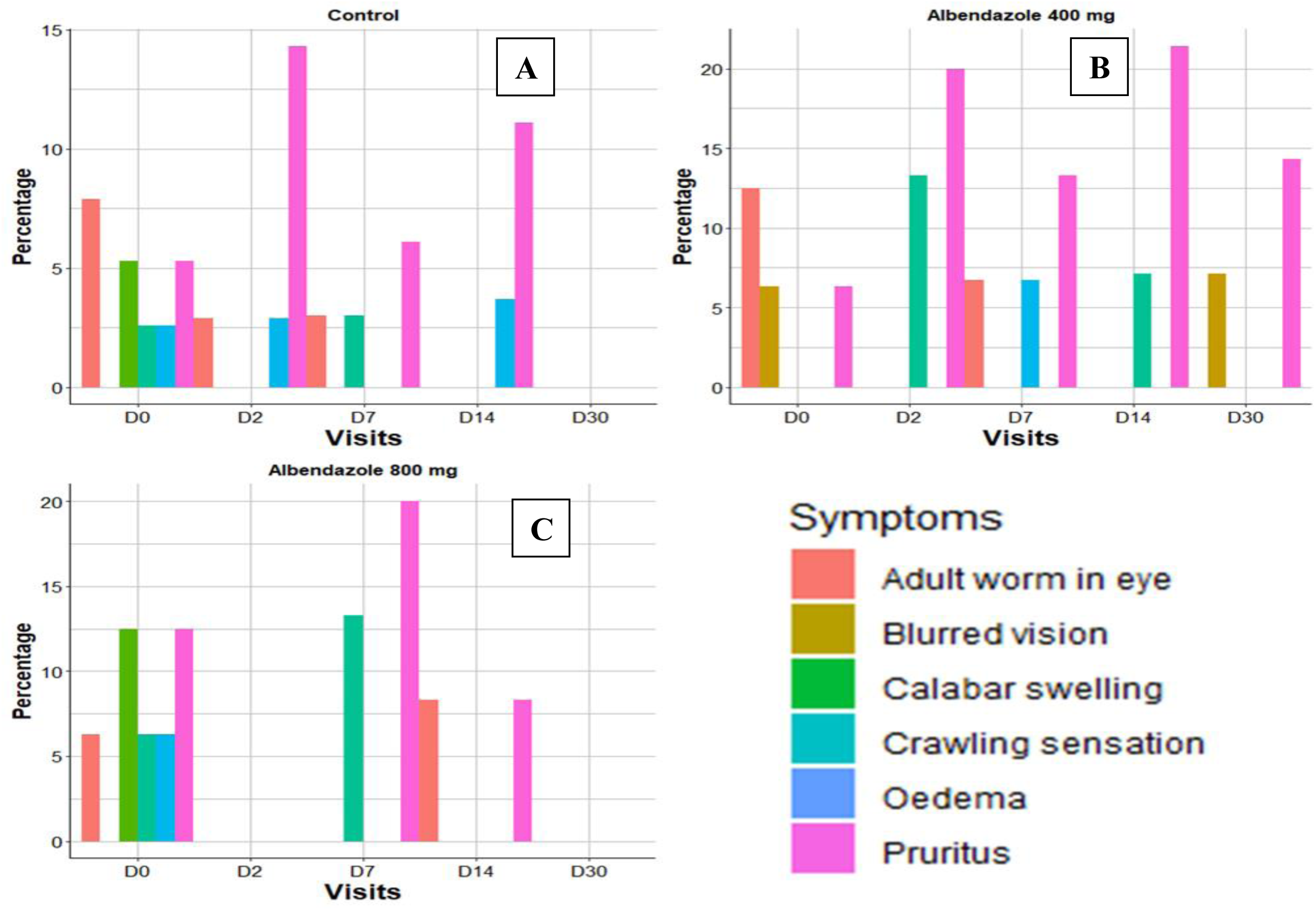
Evolution of symptom frequency from Day 0 to Day 30 in the treatment and control groups.

### Incidence and progression of adverse events during treatment

A total of 11 (31.4%) patients in the control group, 6 (40.0%) patients in the group receiving 400 mg of albendazole, and 4 (28.6%) patients in the group receiving 800 mg of albendazole reported adverse events on Day 2 (*p* = 0.78). By the end of the treatment period, the frequencies of recorded adverse events had decreased, from 31.4% to 0.0% in the control group, from 40.0% to 14.3% in the 400 mg albendazole group, and from 28.6% to 16.7% in the 800 mg albendazole group. However, this decrease was statistically significant only in the control group, where no adverse events were reported after 30 days of treatment (p < 0.01) (Fig 6A).

**Fig 6.**
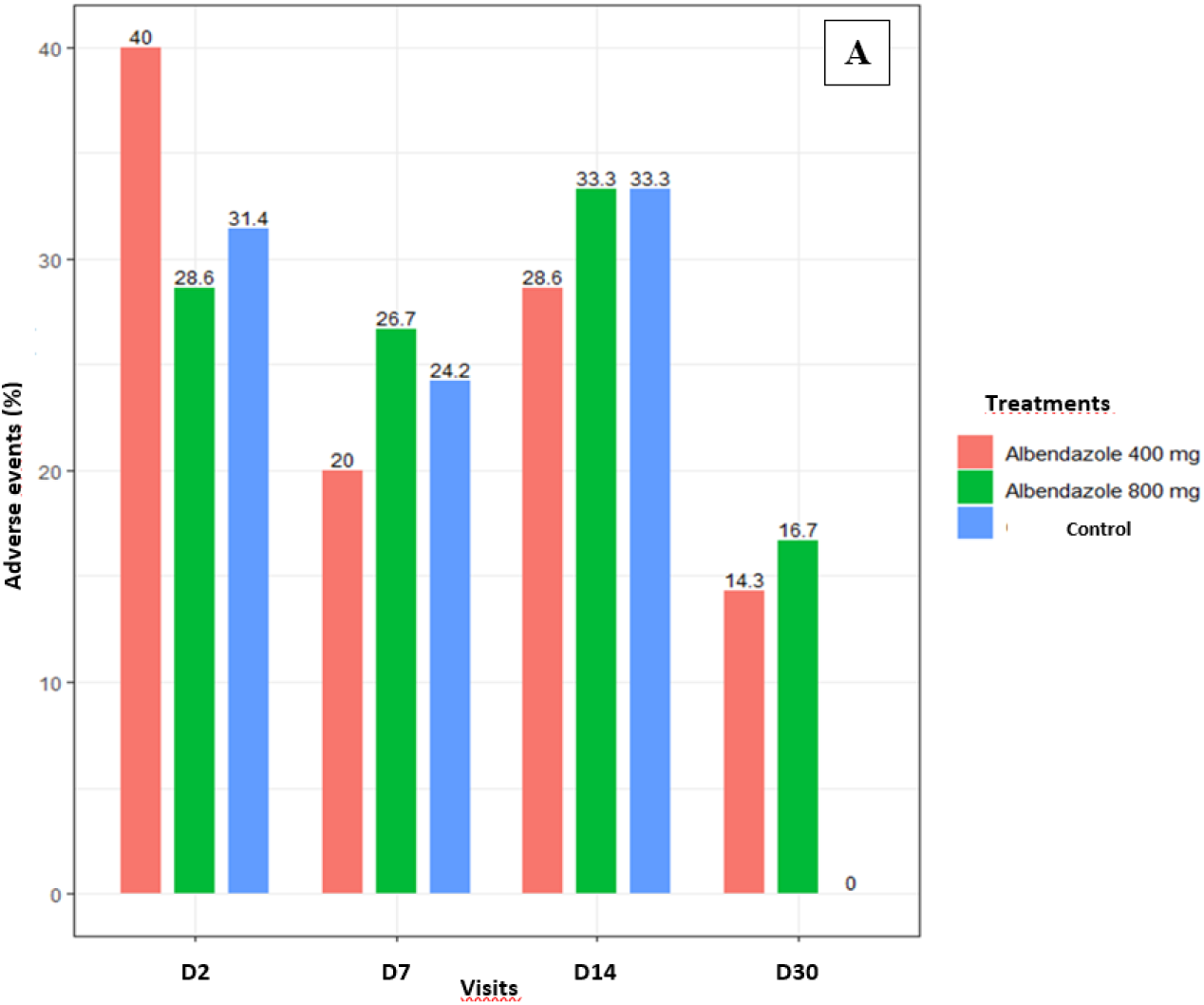
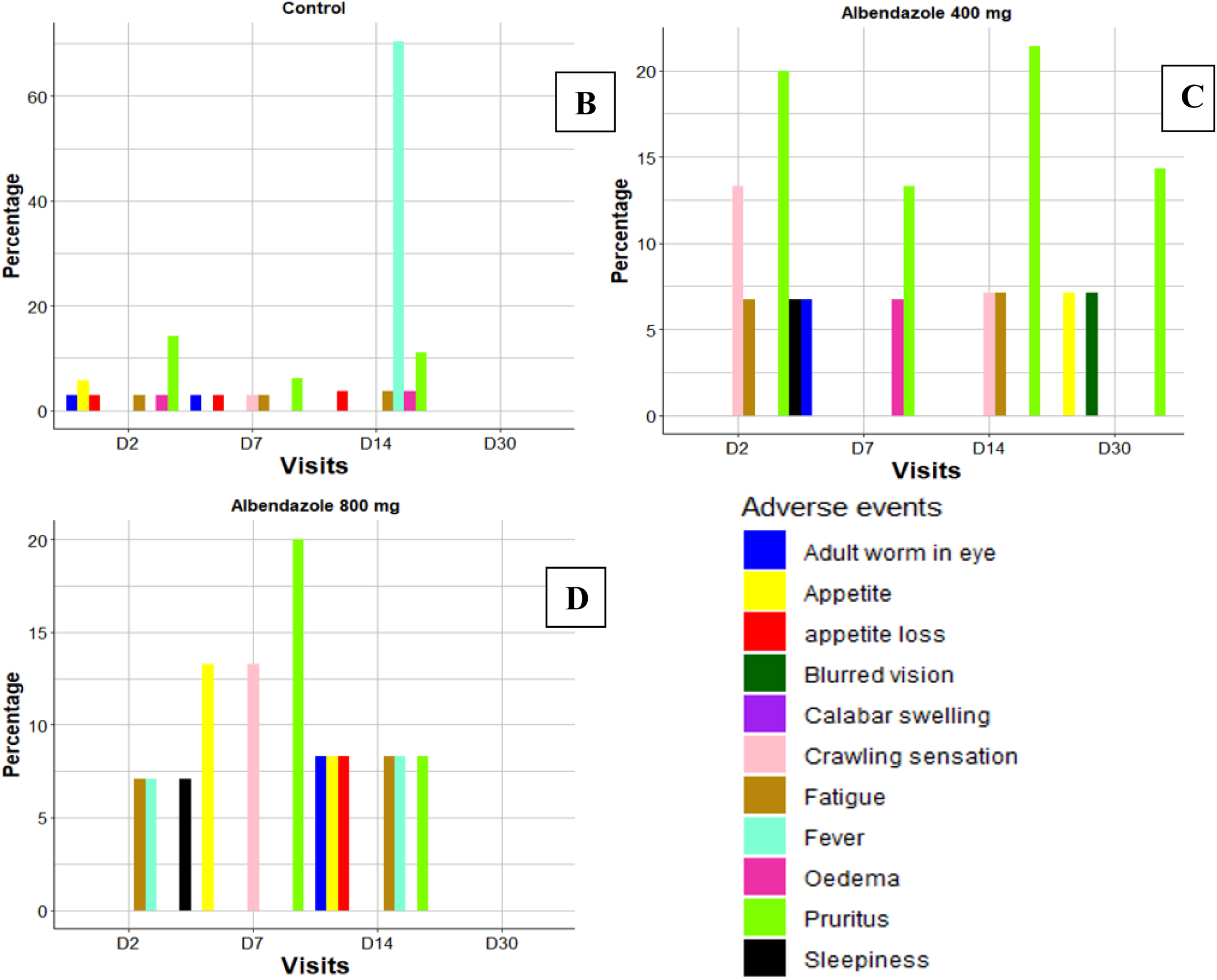
Evolution of adverse event frequency in the treatment and control groups throughout the follow-up period.

On Day 2, in the control group, the most frequently reported adverse event was pruritus, observed in 14.3% of patients. Similarly, in the 400 mg albendazole group, pruritus was also the most common symptom, reported by 20.0% of patients. In the 800 mg albendazole group, asthenia, fever, and drowsiness were the most frequently observed symptoms, each with a frequency of 7.1% (Fig 6A).

On Days 7 and 14, across all treatment groups, although some fluctuations occurred, the overall symptom profiles were comparable. Notably, a high frequency of fever was recorded in 70.4% of participants in the control group on Day 14.

By Day 30, with the exception of the group receiving 400 mg of albendazole—where 14.3%, 7.1%, and 7.1% of patients reported pruritus, increased appetite, and blurred vision respectively—no adverse events were recorded in the other two treatment groups (Fig 6C and 6D).

## Discussion

The Department of Parasitology-Mycology-Tropical Medicine (DPMMT) at the Faculty of Medicine of the Université des Sciences de la Santé has been treating patients with hypermicrofilaraemic loiasis from all regions of Gabon for over 30 years. Elevated microfilaraemia (≥8,000 mf/mL) must be reduced prior to administering diethylcarbamazine (DEC) or ivermectin (IVM). Albendazole (ALB) is the drug of choice due to its low cost and its established use against other helminths, including gastrointestinal and blood parasites such as those responsible for lymphatic filariasis. ALB allows for a gradual reduction in microfilaraemia before initiating macrofilaricidal therapy, facilitating patient recovery [15]. Although therapeutic protocols have been developed, they have not yet been validated using methodologies recognised in experimental drug development within clinical research i.e. methods accepted by the scientific community and necessary for standardisation and deployment in all Loiasis-endemic regions [11,16] Given the co-endemicity of loiasis and onchocerciasis in areas such as Gabon, these protocols could render the entire population in endemic areas eligible for the eradication of this neglected tropical disease. Furthermore, recent studies suggest that hypermicrofilaraemia may be associated with higher mortality rates, particularly among young adults [17,18].

Treatment of loiasis must be effective across different strata of microfilaraemia: low, moderate, and high. We recently reported data from a clinical trial evaluating ALB versus IVM in patients with low microfilaraemia ranging from 500 to 3,500 mf/mL [11]. Another study conducted in Lambaréné assessed patients with microfilaraemia between 5,000 and 50,000 mf/mL treated with ALB over 35 days [19] Additional data from various settings in Gabon are needed to address the critical gap in the adoption of a standardised national protocol, not only for the pre-treatment of hypermicrofilaraemic loiasis but also for the routine management of infected patients.

This randomised, controlled, single-blind Phase IIb clinical trial was conducted to evaluate patients with microfilaraemia above the 8,000 mf/mL threshold. The main findings indicate that hypermicrofilaraemic patients treated with ALB 400 mg and 800 mg for 30 days achieved microfilaraemia reductions below 8,000 mf/mL in 71.4% and 83.3% of cases, respectively, in the intention-to-treat (ITT) analysis, and in 71.4% and 75.0%, respectively, in the per-protocol (PP) analysis. Although the percentage was higher with the 800 mg dose, the difference was not statistically significant. This suggests that increasing the dose may not confer additional benefit in reducing microfilaraemia below the critical threshold of 8,000 mf/mL. Regarding patient adherence, it is preferable to administer a single tablet rather than two, as supported by literature on compliance with chronic disease treatments [20].

To date, no other protocols in the literature match the approach used in this Phase IIb trial. However, some previous studies have investigated similar regimens. For example, Tsague-Dongmo and colleagues developed a treatment based on ALB 800 mg administered over three days for patients with microfilaraemia >8,000 mf/mL. A progressive reduction in microfilaraemia was observed over the first three months, although no significant difference was reported [21]. This supports our findings, indicating that albendazole may induce a gradual decrease in microfilaraemia.

In another study conducted in Gabon, hypermicrofilaraemic patients were treated with ALB 800 mg, administered in two doses of 400 mg over 35 days. This trial reported a significant 91.0% reduction in microfilaraemia (95% CI: 44.0–99.0) after 30 days [19]. Although this reduction was approximately 8% higher than that observed in our study, the difference may be attributed to variations in inclusion criteria, particularly regarding microfilaraemia levels. In that study, the median microfilaraemia was 9,225 mf/mL (7300–16,025), with patients included having levels from 5,000 to 50,000 mf/mL, roughly half of the median parasitaemia detected in our study (17,000 mf/mL [8,200–105,900]), where only patients with >8,000 mf/mL were included [19]. Despite these methodological differences, the findings of Zoleko-Manego and colleagues align with ours, demonstrating an overall downward trend in microfilaraemia. Indeed, in these various trials, a significant reduction in microfilaraemia below 8,000 mf/mL was observed before Day 7 of treatment [19]. This underscores the efficacy of an 800 mg albendazole regimen in reducing microfilarial loads in patients with high parasite burdens, who are at increased risk of severe adverse events if treated with diethylcarbamazine (DEC) or ivermectin (IVM). However, Zoleko-Manego and colleagues did not include a 400 mg albendazole arm. Our results show no significant difference between the 800 mg and 400 mg groups, suggesting that increasing the dose may not confer additional benefit in reducing microfilaraemia below 8,000 mf/mL. These findings may inform optimisation of treatment protocols by balancing efficacy with the drug burden.

Regarding adverse events, 40.0% of patients in the 400 mg group and 28.6% in the 800 mg group reported adverse events after two days of treatment. One might anticipate a higher rate in this study compared to that of Zoleko-Manego and colleagues, given the high median microfilaraemia levels in both populations. Interestingly, the adverse event rates were relatively similar, suggesting that higher microfilarial loads are not necessarily associated with increased frequency or severity of adverse events during albendazole therapy. While the current study, alongside that of Zoleko-Manego et al., supports the safety of albendazole in populations with heavy parasite burdens, there has been at least one report of a serious adverse event following treatment. Métais and colleagues described a case of *L. loa* encephalopathy after albendazole therapy, with loiasis being the only diagnosed pathology after ruling out other causes of coma [22]. This highlights the importance of developing clear, validated protocols for treating loiasis, given the potential risk of cerebral inflammation. Nonetheless, albendazole remains a promising alternative, with the 400 mg once-daily dose appearing optimal.

At baseline, 15.7% of patients presented with at least one symptom. The most frequently observed were ocular migration of the worm (26.7%) and pruritus (24.1%). Over the course of follow-up, clinical symptoms gradually decreased across all groups. After 30 days, no symptoms were reported in either the 800 mg or the control group. However, in the 400 mg group, 14.3% of patients still reported pruritus, and 7.1% experienced blurred vision. Although symptoms persisted in some patients, they were minor, indicating a favourable clinical response. These findings contrast with those of Gobbi and colleagues, in whom 50.0% of patients treated with 400 mg/day of albendazole remained symptomatic after 30 days [23].

Overall, all experimental regimens were found to be effective and well tolerated in this clinical trial. Furthermore, no significant difference was observed between the two intervention groups, despite one receiving double the dose.

This suggests that the 400 mg once-daily regimen is optimal, demonstrating similar efficacy and safety to the 800 mg regimen (administered as two 400 mg tablets). This finding is particularly important as it may reduce drug pressure while maintaining therapeutic efficacy. Additionally, dose reduction could offer clinical and practical advantages. A lower dose may minimise potential side effects in case of very high microfilaraemia and improve adherence, while also reducing treatment costs, potentially enhancing access in rural, low-resource settings.

It should be noted that the study population was relatively older, with a mean age over 45 years across treatment groups. This is likely due to the predominance of older individuals in rural Gabonese villages and the known increase in loiasis prevalence with age in these settings [11]. Socio-economic and environmental factors, such as migration of young adults to urban areas for work or education, may also contribute to this trend.

Our study fits within this context by evaluating the efficacy and safety of albendazole at 400 mg, a drug already widely used for helminthiases, but whose potential in reducing microfilarial loads in hypermicrofilaraemic patients is not yet fully exploited. Providing data on the minimal effective dose contributes to the optimisation of therapeutic protocols, facilitating their deployment in rural resource-limited settings where loiasis control remains a public health priority. The findings of this study have significant public health implications, particularly in establishing an effective and affordable pre-treatment strategy to reduce microfilaraemia prior to administering high-risk treatments such as diethylcarbamazine (DEC) or ivermectin (IVM). The simplicity of administering the 400 mg once-daily regimen could facilitate its large-scale adoption within national loiasis control programmes, especially in rural settings where healthcare access is limited. Furthermore, this approach has the potential to enhance the safety profile of mass drug administration campaigns by decreasing the incidence of severe adverse reactions, thereby increasing prevention coverage, a critical factor in achieving disease elimination.

The main limitation of this study was the absence of follow-up over longer durations to assess the persistence of therapeutic effects, since it is known that complete parasite clearance cannot be observed after only one month of treatment; effective clearance typically occurs at 3 or 6 months with additional molecules.

For a Phase IIb clinical trial, the relationship between microfilaraemia and albendazole metabolite concentrations (pharmacokinetics) should also be evaluated. Furthermore, the DPMMT home-protocol has demonstrated full patient recovery at 6 months (Moutombi 2025, submitted).

Further studies are needed to confirm the long-term sustainability of the therapeutic effects, including follow-up over 6 to 12 months. The pharmacokinetics of albendazole in this specific continuously-exposed population should also be better characterised to optimise dosing and minimise the risk of rare adverse events, particularly neurological complications. Additionally, reproducibility of these results across different epidemiological and geographic contexts should be evaluated to facilitate wider implementation.

## Conclusion

This study demonstrates that albendazole at 400 mg once daily for 30 days is as effective and well tolerated as the 800 mg regimen in patients with hypermicrofilaraemia. The comparable efficacy, combined with the potential benefits of lower drug burden, reduced costs, and improved adherence, supports the adoption of the 400 mg regimen as the optimal protocol in endemic settings while limiting potential adverse effects. These findings contribute valuable evidence towards the development of standardised, validated pre-treatment strategies that can safely reduce microfilarial loads, facilitating the safe administration of ivermectin for MDA or diethylcarbamazine for treatment. Implementing such protocols could significantly advance efforts to control and eliminate loiasis in endemic regions, ultimately reducing disease burden and associated morbidity.

## Data Availability

All data underlying the findings described are within the manuscript itself.

## Acknowledgments

The authors are grateful to all the staff at the Centre de Recherche biomédicale En pathogènes Infectieux et Pathologies Associées (CREIPA) and the Unité Mixte de Recherche sur les Agents Infectieux et leur Pathologie (UMRAIP) for their support in participant recruitment in Bitam, and for their assistance with sample processing in both Libreville and Bitam. Special thanks are also to Bifolossi Medical Center staff for hosting our study. We extend our gratitude to the village chiefs and other relevant authorities for their support throughout this research. Espediee CHABI, Junior Dimitri MOUDOUMBI-MOUANDZA, Stéphane OGOULA, Ivan-Guy-Tiburce NDAMA-NDAMA, Lys Anna HOMBO NDOMANA, Melly Chancelle MIMBILA MINKO, Ndjena Andress OBALI, Télesphore OBIANG NGOUA and Rochat Léotard SIMA OWONO for their work on the field and in the laboratory. Finally, we thank all the participants whose cooperation made this study possible.

## Author contributions

## Conceptualization

Marielle Karine Bouyou Akotet, Noé Patrick M’Bondoukwé

## Data curation

Luccheri Ndong Akomezoghe, Luice Aurtin Joel James, Noé Patrick M’Bondoukwé,

## Formal analysis

Luccheri Ndong Akomezoghe, Ginette Sévérine Zang Ondo, Luice Aurtin Joel James, Noé Patrick M’Bondoukwé

## Funding acquisition

Marielle Karine Bouyou Akotet, Noé Patrick M’Bondoukwé

## Investigation

Luccheri Ndong Akomezoghe, Hadry Roger Sibi Matotou, Bridy Chesli Moutombi Ditombi, Coella Joyce Mihindou, Valentin Migueba, Jacques Mari Ndong Ngomo, Noé Patrick M’Bondoukwé, Bedrich Pongui Ngondza, Christian Mayandza,

## Methodology

Luccheri Ndong Akomezoghe, Denise Patricia Mawili-Mboumba, Noé Patrick M’Bondoukwé

## Project administration

Denise Patricia Mawili-Mboumba, Marielle Karine Bouyou Akotet, Dimitri Hardrin Moussavou Mabicka, Héléna Kono

## Resources

Marielle Karine Bouyou Akotet

## Supervision

Denise Patricia Mawili-Mboumba, Marielle Karine Bouyou Akotet

## Validation

Denise Patricia Mawili-Mboumba, Marielle Karine Bouyou Akotet, Charleine Manomba

## Visualization

Luccheri Ndong Akomezoghe, Luice Aurtin Joel James, Noé Patrick M’Bondoukwé

## Writing-original draft

Luccheri Ndong Akomezoghe, Ginette Sévérine Zang Ondo, Noé Patrick M’Bondoukwé

## Writing-review and editing

Bridy Chesli Moutombi Ditombi, Marielle Karine Bouyou Akotet, Denise Patricia Mawili-Mboumba

